# Multiple Treatment Interruptions and Protecting HIV-Specific CD4 T-Cells Enables Durable CTL Response and Viral Control

**DOI:** 10.1101/2023.10.24.23297421

**Authors:** Anshika Jain, Gaspar E. Canepa, Mei-Ling Liou, Emily L. Fledderman, Andrei I. Chapoval, Lingzhi Xiao, Ipsita Mukherjee, Jeffrey A. Galvin, Princy N. Kumar, José Bordon, Marcus A. Conant, Jefferey S. Boyle

## Abstract

**BACKGROUND:** The cell and gene therapy product AGT103-T was evaluated (NCT04561258) for safety, immunogenicity, and persistence for up to 180 days post infusion. We sought to investigate the impact following analytical treatment interruptions (ATIs).

**METHODS:** Six patients suspended their antiretroviral therapy (ART) until their viral load reached 100,000 copies/mL in two successive visits, or their CD4 count fell below 300 cells/μL. We measured the magnitude of viral rebound, the persistence of AGT103-T transduced CD4+ T-cells and the impact on HIV-specific immune responses.

**RESULTS:** During the ATI, all patients experienced logarithmic viral rebound followed by a 2-5-fold increase in total CD8 counts, that coincided with a rise in HIV-specific CD8 T-cells. This was attributed to the increase in antigen availability and memory recall. Thus, to determine if the immune response generated during this “auto-vaccination” event can contribute to viral suppression upon subsequent exposures, a second ATI was initiated. During the second ATI, the Gag-specific CD8 T cells were either maintained or rose and the peak viremia was substantially decreased with viral set-points ranging from 7,000-25,000 copies/mL. Upon ART resumption, faster viral control was demonstrated without any serious adverse events (SAEs) or drug resistance.

**CONCLUSION:** AGT103-T gene therapy and multiple ATIs were not associated with SAEs and allowed subjects to establish a low viral set-point with relatively stable CD4 T cell counts. Additionally, multiple ATIs are beneficial for the study design when induction of CD8 T cells is required to establish viral control.

**REGISTRATION:** ClinicalTrials.gov NCT05540964

**FUNDING:** American Gene Technologies International Inc.

## INTRODUCTION

Despite the efficacy of highly active antiretroviral therapy (HAART), which allows for a durable long-term remission from Human Immunodeficiency Virus-1 (HIV-1), the need for the life-long, strict adherence, long-term associated toxicities, potential development of antiretroviral (ARV) drug resistance strains, and its inability to eradicate the latent virus necessitate further research towards finding an HIV cure. This will likely require innovative and multi-faceted strategies. Several studies have indicated that a preserved or augmented virus-specific cytotoxic T-lymphocyte (CTL) response in the presence of CD4 helper T-cells is vital for sustained virologic control (1, 2). During the primary infection, while a robust virus-specific CD4-mediated CTL response associated with partial control in viremia is generated, due to the diversification of virus around the peak of the infection and limited functionality of the CTL response, the immune system is unable to clear the virus (3). Furthermore, HIV specific CD4 T-cells that are critical for maintaining an effective and sustained anti-HIV immune response, are preferential targets of HIV infection (4, 5), and are therefore additionally targeted by the CTL response. This is further complicated by the integration of the viral genome with the host cell DNA, resulting in continuous infectious cycles leading to a progressive loss of CD4 T cells. Thus, the declining CD4 T cells, combined with persistent presence of the antigen, leads to T cell exhaustion and the failure to generate an effective memory response (6). Overall, this interplay between the virus and the immune system complicates the path to eradicating HIV. If left untreated, a stable level of viremia, referred to as the viral set-point, is attained after the resolution of primary infection. This set-point is governed by both ongoing viral replication as well as exhausted immune responses, which persists for several years before cellular immune responses are completely exhausted, and the patient progresses towards AIDS.

Although HAART can result in the restoration of total CD4 counts, the HIV-specific CD4 T cells are not fully restored (7). Therefore, upon cessation of ART, only a modest CTL response is generated, which is ineffective in mediating viral suppression or viral clearance (8). Thus, strategies to protect the CD4 T cells from new infections that can elicit a sustained CTL response in the presence of viremia are vital for achieving a functional cure against HIV. In addition to insufficient levels of HIV-specific CD4 helper cells in HIV patients, another major obstacle to viral clearance is the presence of latent viral reservoir in immune cells. While actively transcribing cells can be detected by the immune system, due to limited transcription of the integrated pro-virus, latently infected cells cannot be targeted by immune effector mechanisms and present themselves as a source of viremia that can result in further depletion of the CD4 T cells (9). However, the efforts in the field of immune, gene and cell therapies have provided encouraging data that suggests that enhancing the immune response of patients while suppressing HIV genes has a promising shot towards eradicating HIV.

To infect the CD4 T cells, HIV relies on binding to the CD4 receptor and at least one of the two co-receptors, CCR5 and CXCR4. CCR5-tropic strains are predominant during the early stages of infection, while CXCR4 tropism may arise with disease progression (10). This sequential binding results in the fusion of HIV and host membranes, thus initiating the process of infection. The remission from HIV in the “Berlin patient”, attributed to a natural Δ32 homozygous mutation in the *CCR5* gene that resulted in a non-functional co-receptor, spawned the idea of *CCR5* inhibition as a potential therapeutic mechanism against HIV infection. Employing this strategy, adoptive T-cell therapies have been developed, wherein autologous CD4 T cells from HIV+ patients have been modified ex-vivo to inhibit the expression of *CCR5* and are then infused back into the patients (11–13). Among the other strategies devised to protect infused CD4 T cells from HIV, include overexpression of restriction factors, use of fusion inhibitors, targeting HIV coreceptors through RNA and gene editing techniques, targeting HIV genes through siRNA, dominant negative genes, and antisense RNA, only a handful have made into the clinic (13). A clinical trial with an autologous T-cell product that is modified with zinc finger nuclease-mediated *CCR5* suppression demonstrated a modest clinically significant response upon cessation of ART, showing a delayed viral rebound associated with an improved CD8 T cell response in a small subset of the population, with one subject demonstrating immunologic viral escape (14). Despite the encouraging data demonstrating a sustained immune response, the long-term therapeutic persistence of these cells and their role in viral suppression has been challenging with current clinical therapies. This challenge may be attributed to the sub-optimal/sub-therapeutic level of HIV-specific cells within these drug products, which is insufficient to generate an effective immune response leading to a significant viral suppression.

Given the remarkable virologic suppression achievable by ART, analytical treatment interruption (ATI) is likely to remain necessary for assessing the therapeutic efficacy of HIV treatments. Multiple studies have established that short bursts in viremia during ATI do not result in an expansion in the persistent viral reservoir, nor cause an irreparable immune dysregulation, justifying the inclusion of ATI in evaluating HIV curative strategies (8, 15). Multiple ATIs or repeated exposures to such short bursts of viremia have been proposed to boost immunity, a theory referred to as “auto-vaccination” (16, 17). However, its correlation with overall enhanced immunogenicity and viral suppression remains controversial. While some reports with a small number of participants have suggested its beneficial effect in increasing the breadth of CTL response and viral suppression (2, 18), studies conducted on larger cohorts have not found the association between the amplified CD8 response and a viral suppression (16, 17, 19, 20). Nevertheless, no harmful effects of multiple ATIs were reported in any study. Moreover, the effect of multiple ATIs and induction of CD8 T cell responses in the setting of cell and gene therapy has never been investigated.

To achieve a functional cure, we developed a strategy that constitutes mechanisms that protect CD4 T cells from infection to enable robust immune responses and thereby reducing the viral burden. We developed an autologous CD4 T cell product, AGT103-T, that comprises a highly enriched population of Gag-specific CD4 T cells, essential for generating the CD8 T cell response in the presence of viremia (12, 21). Furthermore, these cells were genetically engineered using a 3^rd^ generation, self-inactivating lentiviral vector, AGT103. This vector expresses a cluster of three microRNAs targeted towards inhibiting human-*CCR5*, the transcripts encoding for *HIV-Vif*, and *HIV-Tat* under a constitutive promoter. The integration of this cassette not only results in protecting the cells from new infection (by inhibiting *CCR5* expression) but also inhibits viral transcription by targeting two essential HIV genes, *Vif* and *Tat*, thereby disabling virion formation and reducing the overall viral reservoir. Given that Gag-specific cells are preferentially infected by HIV (4) and transduced by lentiviral vector following the in vitro stimulation (21), our drug product provides HIV-specific cells that are resistant to HIV infection and therefore, can potentially and durably withstand viral rebound whilst contributing to enable an effective CTL response. In our previous clinical trial (NCT04561258), we demonstrated that AGT103-T was safe for human administration and was well tolerated by all seven participants without any Grade 3 or higher adverse events reported (12). Among the seven participants enrolled in the study, five participants (02-001, 01-002, 01-005, 01-006, 01-007) were infused with a low dose: ≥1 × 10^8^ and < 1 × 10^9^ of AGT103-transduced CD4 T cells and two (01-008 and 02-009) were infused with a high dose: ≥1 × 10^9^ and < 5 × 10^9^ AGT103-transduced CD4 T cells. The cell product also contained an enriched population of Gag-specific cells that constituted up to 30% of the total viable cells. Although the proportion of cells that were Gag-specific as well as AGT103+ was not determined in the clinical product, approximately 50% of cells were shown to be double positive in our pre-clinical model (21). Upon infusion of the clinical product, a dose-dependent engraftment of AGT103-T cells was observed in peripheral blood, that peaked 3 days post-infusion (12). Similarly, 7.5 to 300-fold expansion of the Gag-specific CD4 T cell response that peaked around 14 days post-infusion was also observed in all participants (12).

To assess the impact of AGT103-T on the viral rebound, HIV immune responses, patient safety, and to prevent the loss of critical information if subjects ceased their ART without medical supervision (as was observed in the case of two study participants), we developed an ATI study, NCT05540964. Based on the initial clinical data obtained from the study and in the light of previously reported effects of “auto-vaccination” on HIV immunity, we hypothesized that two sequential ATIs in the presence of HIV-resistant Gag-specific AGT103-T cells could substantially boost the CTL response and lead to significant viral suppression. Herein, we present the results of our clinical trial demonstrating the relevance and benefits of sequential ATIs in eliciting and measuring the efficacy of cell and gene therapy against HIV. We show that multiple ATIs can contribute to generating an amplified CD8 T cell response. Overall, our findings demonstrate that multiple ATIs after AGT103-T treatment can result in significant viral suppression. The lower viral set-points established after AGT103-T enable faster control with ART and may be useful in improving regimens in the future.

## RESULTS

### AGT103-T Treated Participants Demonstrated an Amplified CD8 T Cell Count After the First ATI

The study participants initiated the first ATI 99 to 490 days post-infusion, and their viremia along with other immune parameters were measured weekly for a month and monthly thereafter. Consistent with the previous studies including ATI (22), a rapid viral rebound was observed in all participants upon cessation of ART, with the observed peak at a median viral load (VL) of Log_10_ 5.4 copies/mL (Range: Log_10_ 3.429 to Log_10_ 5.76 copies/mL) attained between 30-56 days (Figure 1). Notably, participant 01-007, who started HAART during early phase of infection, achieved viral control with undetectable levels of viremia for 91 days upon cessation of drugs (based on patient communicated timeline and data), with a transient blip to 2690 copies/mL that spontaneously decreased to 80 copies/mL during the ATI (Figure 1A). Overall, this participant exhibited a viral control under 3000 copies/mL for approximately six months. All participants resumed ART post-viral rebound at different times. 50% of the participants (02-009, 02-001 and 01-002; Figures 1B, 1C, 1E) met the ART resumption criteria within 30-54 days, the others (01-005, 01-007, and 01-008; Figures 1D, 1A, 1F) resumed ART without meeting the resumption criteria. Participant 01-005 reached their first VL >100,000 copies/mL 56 days after the initiation of ATI, which spontaneously declined to 69,200 copies/mL at their next monthly visit (day 77) (Figure 1D). Despite the downward trend in viremia, resuming ART without meeting the criteria was recommended to prevent further viral reservoir accumulation or prolonged virus exposure. Participants 01-007 and 01-008, who self-initiated their ATI before enrolling in the study also started ART without meeting the criteria. Patient 01-008 deviated from the protocol during the safety study (NCT04561258), self-initiated an ATI, and exhibited a VL of 570,000 copies/mL in the monthly visit, prompting immediate ART resumption (Figure 1F). Patient 01-007 resumed ART with a VL <100 copies/mL and withdrew from the study for personal reasons after the first ATI.

**Figure 1:**
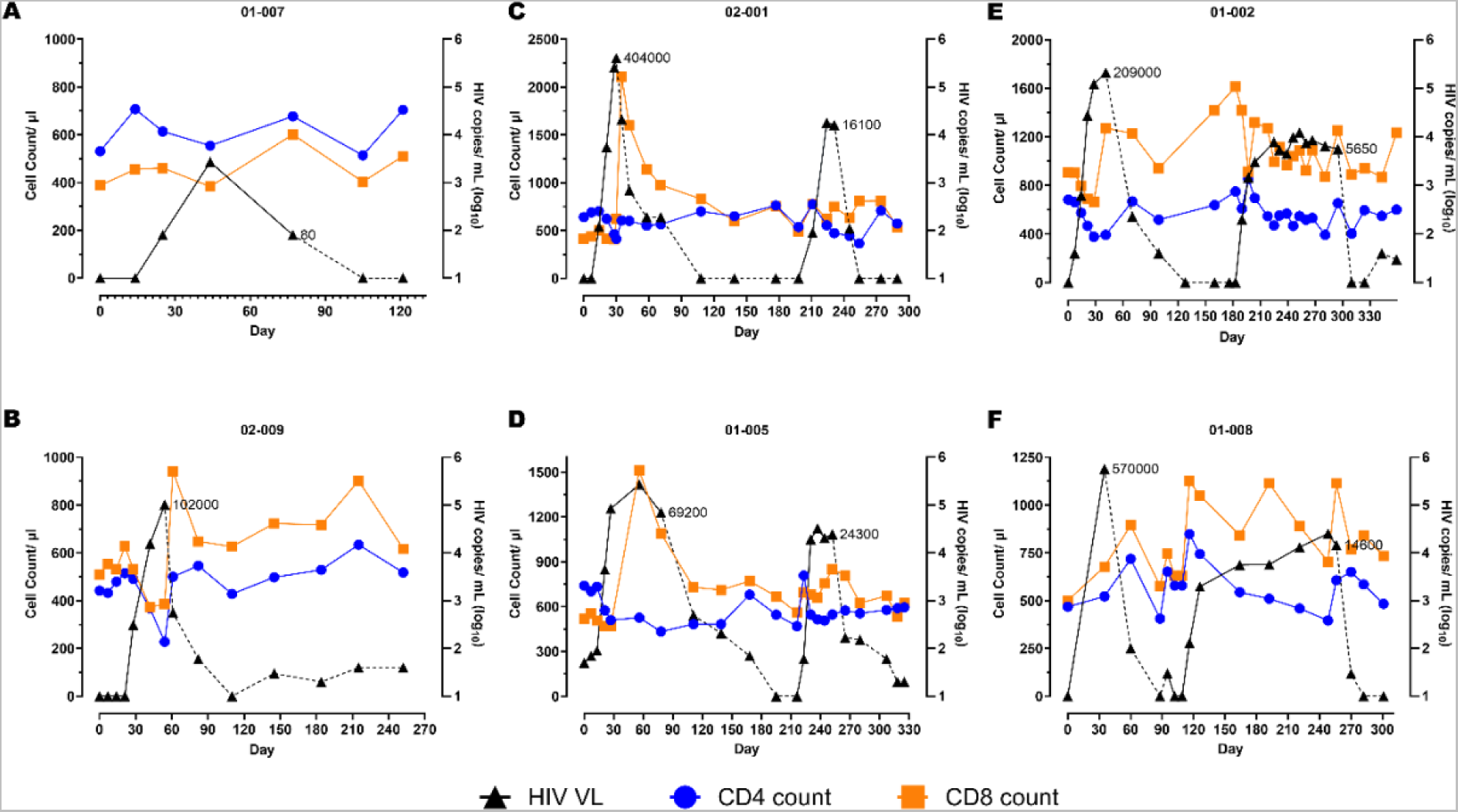
Two Analytical Treatment Interruptions (ATIs) Enable Viral Suppression in AGT103-T Treated Participants. (A-F) These figures illustrate the impact of (A, B) one ATI or (C, D, E, F) two ATIs on viral rebound and immune parameters (total CD4 and CD8 T cell counts) in six participants. The participant IDs are indicated above each figure. Participants who were previously infused with AGT103-T, underwent one or two ATIs 99-490 days after the infusion. Day 0 is referred to as the first day of the ATI after ART cessation. Viral load (VL) is represented on the right-hand Y-axis and total CD4 and CD8 T cell counts are represented on the left-hand Y-axis as measured at the indicated timepoints. Participant 01-008 self-initiated his first ATI during the previous safety study. Participant 01-007 self-ceased their ART 77 days prior to their enrollment and exhibited viral control for a total of 91 days (pre-enrollment data is not shown here). The VL (HIV copies/mL) at which the participants resumed ART are indicated along the viral load curve in each graph. HIV viral copies measured in plasma are represented by black triangles (copies/mL Log_10_), solid black line corresponds to ATI, dashed black line corresponds to ART; blue circles represent CD4 cells (count / mL of blood); orange squares represent CD8 cell (counts/mL of blood).

We then measured the immune parameters and their correlation with the rise in viremia. Upon the viral rebound, the median circulating CD4 T cell count declined from 585.5 cells/µL (range: 442 to 740 cells/µL) to 410 cells/µL (range: 228 to 514 cells/µL) with only one participant, 02-009, whose CD4 T cell counts decreased below 300 cells/µL leading to recommendation of ART resumption (Figure 1). Upon re-institution of ART, CD4 T cell counts in all participants returned to baseline levels or within 15% of the baseline. On the other hand, a notable increase in total CD8 T cell counts, peaking 2 to 5-fold above baseline, following the viral peak was recorded in all participants. The median CD8 T cell counts of 504.5 cells/µL (range: 388 to 907 cells/µL) at baseline increased to 1107 cells/µL (range: 601 to 2106 cells/µL) following the peak in viral rebound (Figure 1). This response is comparable to that observed in a subset of participants in another study involving the infusion of autologous CD4 T cells with *CCR5* knockdown (14) but contrasted with the CD8 counts reported in other ATI studies without any therapy where a decrease in CD8 count has been observed upon ATI (8). The amplified response seen in all 6/6 of our patients suggests a role for AGT103-T in enhancing systemic CD8 T cell counts. Upon ART resumption, a modest decrease in CD8 T cell counts was observed, however, their overall levels remained above baseline in all participants (Figure 1).

With the assumption of the first ATI serving as an “auto-vaccination” event (2, 17, 18, 20) and based on the amplified CTL count observed following the viral rebound in all the study subjects, we hypothesized that this will allow the generation of an effective immunogenic response leading to viral suppression upon a subsequent viral exposure. Consequently, we offered a second ATI to all our study participants.

### Augmented CD8 T Cell Counts and Lower Viral Rebound was Observed During the Second ATI

Upon the start of the second ATI, three of the four participants experienced a similar or delayed viral rebound (Figure 1C-1F). Notably, patient 01-005, who had a longer duration of 78 days in the first ATI, experienced a more rapid viral rebound, which is likely due to their increased viral reservoir resulting from the longer duration in the first ATI (Figure 1D). Although the magnitude of the viral rebound was variable, all participants demonstrated a significant viral suppression compared to the first ATI. During the second ATI, a median peak viremia of 4.3 Log_10_ copies/mL (Peak VL range: Log_10_ 4.1 to Log_10_ 4.4 copies/mL), which was 1.2 Log_10_ lower than the median peak viremia (Log_10_ 5.52 copies/mL) during the first ATI. All participants attained their set-point (median 4.13 Log_10_ copies/mL; range= 3.86 to 4.38 Log_10_ copies/mL), within 1-2 weeks (Figures 1C-1F). These observations are interesting because a significant viral suppression from multiple ATIs has not yet been demonstrated (2, 17). Additionally, a viral set-point, that is typically achieved 4-6 weeks after the peak viremia (23), was attained much earlier after the second ATI in our study participants. While 02-001 (Figure 1C) and 01-005 (Figure 1D) resumed their ART 35 days post-ATI, 01-002 (Figure 1E) and 01-008 (Figure 1F) were able to maintain their VL set-point at approximately 7,560 (Log_10_ 3.86) copies/mL and 12,894 (Log_10_ 4.04) copies/mL, respectively for over 120 days. Furthermore, all participants were able to return to undetectable levels of viremia without developing drug resistance or experiencing viral escape, indicating a positive safety profile. No serious adverse events, or event of viral transmission were recorded after the ATIs.

In addition to the enhanced viral suppression, all the participants demonstrated improved immune parameters during the second ATI. Contrary to a decline observed at the onset of the first ATI, a concomitant increase in both CD4 and CD8 T cell count was observed during the second ATI (Figures 1C-1F). The initial increase in CD4 T cells in response to viral rebound appears to have helped in generating a concurrent and approximately 2-fold amplified CD8 T cell count, that in turn led to significant viral suppression. With the further increase in viremia as the ATI continued, although the total CD4 T cells underwent a gradual decline, the systemic CD8 T cells were able to maintain their levels above baseline (Figures 1C-1F). It is noteworthy that while all the other participants observed an approximately 35% depletion in their CD4 T cells (Figures 1C, 1E, 1F), 01-005 did not observe any decline compared to their CD4 value at the start of the second ATI (Figure 1D). With the resumption of HAART, the CD4 T cells were restored, and CD8 T cells declined (albeit remaining above baseline). Taken together, this data suggests that a short primary exposure to the virus is beneficial to prime the immune system such that it can generate an immune response that can contribute to viral suppression upon subsequent viral exposures.

### An Increased Gag-specific Immune Response is Observed After ATI

To investigate if the immune responses observed were functional against HIV, we determined the HIV-specific immune response by measuring the Gag-specific CD4 and CD8 T cells by intracellular cytokine staining (ICS) assay. Overall, Gag-specific T cells demonstrated a similar trend as observed for the total CD4 and CD8 T cells. An increase in Gag-specific CD8 T cells with the rise in viremia was observed during both ATIs (Figure 2). The magnitude of the CD8 T cell response to Gag peptide was higher during the first ATI (1.1-13-fold) than the second (1.5-5.3-fold), with 01-008 being an exception where a higher Gag specific CD8 response was recorded during the second ATI (1.5-fold higher during first ATI and 5.3-fold higher during the second ATI (Figure 2). Gag-specific CD4 T cells demonstrated higher stability and durability than the total CD4 T cells at the onset and during the presence of viremia. After the first ATI, following the event of “auto-vaccination”, 3 out of 4 subjects (except 01-008) demonstrated an increase in Gag specific CD4 T cell response during the second ATI, although a similar increase in the Gag-specific CD8 response was not generated. Overall, our data indicates that two ATIs helped in increasing the HIV specific CD4 response that led to the CTL response that in-turn coincided with reduced viral load. Given that increase in Gag-specific cells were predominantly increased after the second ATI, our data also suggests that multiple ATIs are essential to boost immunity in the presence of cell and gene therapy products.

**Figure 2:**
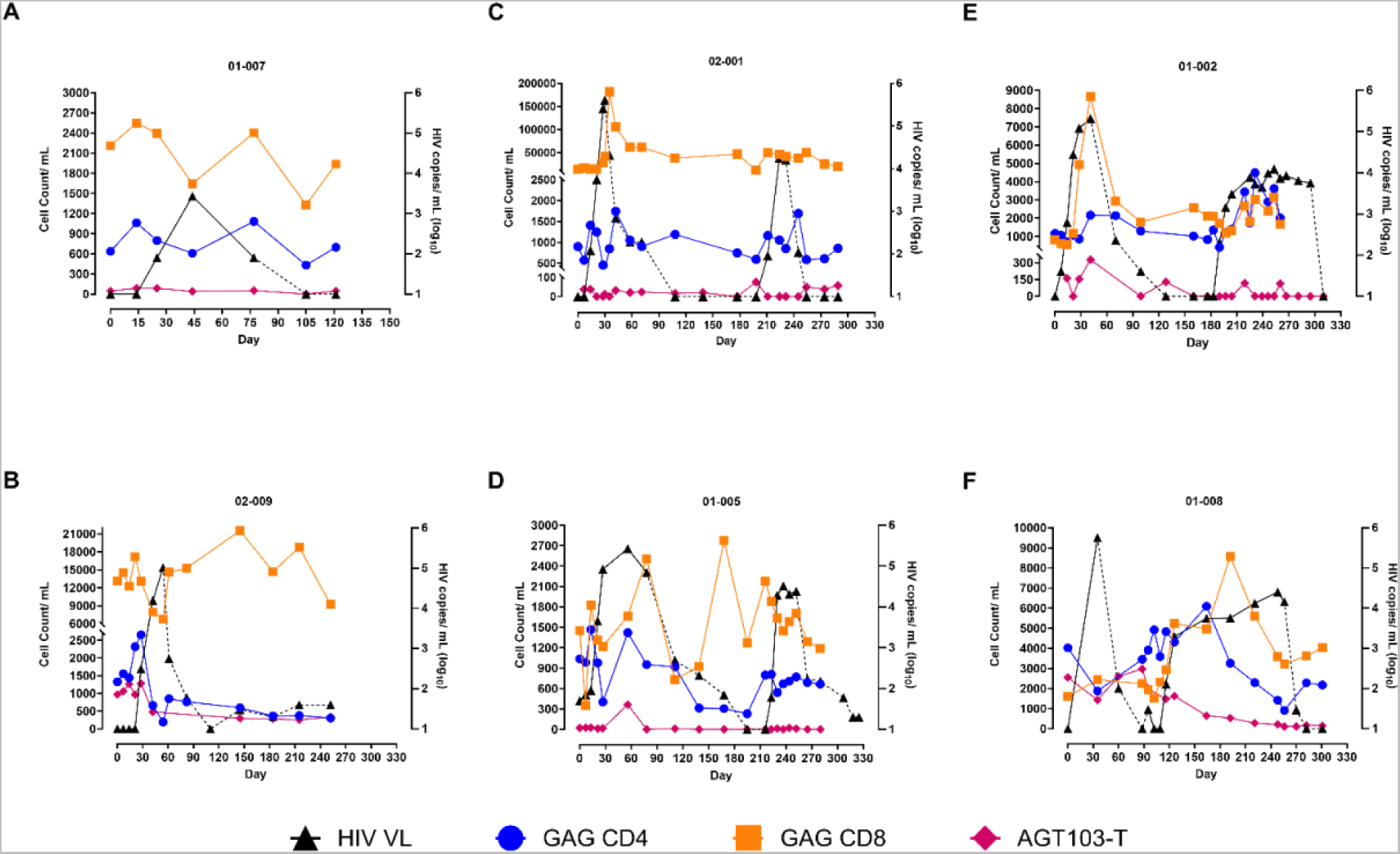
Persistence of AGT103-T and HIV Gag-Specific CD4- and CD8-T cells in Circulation. (A-F) Counts of AGT103-T cells, Gag-specific CD4 and Gag-specific CD8 T cells (left-y-axis) detected per mL of blood at various time points (x-axis) along with the viral load (right-y-axis) are plotted for each of the six participants. The participant IDs are indicated above each figure. The modified cells were assessed by measuring the number of copies of the transgene in cells encoding WPRE, a sequence unique to the lentivirus transduced in the modified cells. HIV-specific immune response was counted by measuring the Gag-specific CD4 and CD8 T cells by intracellular cytokine staining (ICS) assay in six participants. Black triangles correspond to the HIV viral copies in plasma (copies /mL Log_10_); solid black line corresponds to ATI; dashed black line corresponds to ART; blue circles represent Gag-specific CD4 cell count/mL of blood; orange squares represent Gag-specific CD8 cell count/mL of blood; pink diamonds represent AGT103-T cells.

### Persistence of AGT103-T in Circulation

To determine if AGT103-T cells could have contributed to the viral suppression observed during the second ATI, we investigated the presence of AGT103-modified cells in patient’s circulation by two independent methods. These methods detected different sequences of the lentiviral vector. First, we employed a qPCR-based approach to ascertain the vector copy number (VCN) in an enriched CD4 T cell population, identifying cells containing a unique sequence, the woodchuck hepatitis virus post-transcriptional regulatory element (WPRE), present in our lentivirus within the modified cells, normalized to the housekeeping gene *RPL32* (24). Consistent with previously reported dose-dependent engraftment (12), the persisting levels of the modified cells were found to be correlated with the dose and the time since the infusion. A significant proportion of circulating CD4 T cells was detected in the two high dose patients, 01-008 (0.54%) and 02-009 (0.22%) at the beginning of the ATI (Table S1), that were comparable to the number of Gag-specific CD4 cells (Figures 2F, 2B), suggesting a possibility that a significant proportion of Gag-specific cells could be AGT103-modified. Conversely, AGT103+ cells constituted a lower proportion of circulating CD4 T cells in other participants including 01-007, 02-001, 01-002 and 01-005, which can be attributed to their low dose of infusion and a longer duration of 246-490 days post-infusion (Figures 2A, 2C, 2D, 2E, Table S1). A decline in AGT103-T cells occurred with the rise in viremia, which appeared to partially restore upon the resumption of ART, in all participants (Figure 2). This suggests the possibility of migration of AGT103-T cells out of the circulation into the site of viral replication during viral rebound and back into circulation during ART, although the possibility of cell death and proliferation cannot be ruled out. We confirmed these results using the amplicon-sequencing approach and observed a similar pattern of AGT103-T persistence. Since our silencing approach utilizes naturally occurring miR30, we amplified and sequenced miR30 from the DNA obtained from the CD4 T cells and obtained the ratio of modified to unmodified (native) miR30 sequences, that was then used to determine the AGT103+ cells (Figure S1). We also performed the VCN assay on the non-target cells obtained in the flow through (FT) while enriching the CD4 T cells. The absence of WPRE signal from the FT in all the patients suggests that only CD4 T cells were transduced with AGT103 lentiviral vector during drug product manufacture (data not shown). In the event of any rare chance of non-target cell transduction, our data shows that the non-target cells did not persist in the patients that otherwise maintained a significant population of AGT103+ CD4 T cells. This data suggests that enriching a target population of cells can increase the sensitivity of the downstream qPCR-based assays in clinical samples, which is particularly important for detecting very low persisting levels of CAR-T or other transduced cells in circulation that do not express extracellular surface markers. Taken together, the in vivo persistence of AGT103-T cells two years after infusion, surpassing the median lifespan of CD4 T cells, and their fluctuating levels in the presence of viremia indicates their long-term engraftment in the patients that is responsive to the viral rebound and immunological signals. Furthermore, the viral suppression and amplified immune response observed in all our study participants indicate the contribution of our therapeutic product to the immunological enhancement of our study participants, however a direct correlation cannot be established.

### AGT103-T Treatment Causes Viral Load Reduction and Faster Control of Viremia with ART

All four study participants demonstrated lower peaks of viremia after the second ATI. Based on the historical data provided by one of the study participants, 01-005 prior to AGT103-T treatment had an extremely high VL of >10,000,000 copies/mL, above the level of quantitation at time of diagnosis in 2010, which took 384 days on treatment to reach undetectable levels (data not shown). This patient received an infusion of AGT103-T (4.6 X10^8^ transduced cells) in August of 2021, however, did not initiate ATI until 411 days post-infusion. Nevertheless, upon ART withdrawal post-therapy, the patient demonstrated a peak VL of 267,000 copies/mL, which is a 30-fold reduction from the initial diagnosis. Subsequently, they reached a VL of 69,200 copies/mL, 140-fold reduction from their original peak viremia at diagnosis. During the second ATI, this patient observed a peak viral rebound to 24,300 copies/mL that represents over 400-fold reduction in VL from the initial diagnosis. Along with a significant decrease in the viral rebound after each ATI, this patient also observed a remarkable decrease in the number of days required to reach the undetectable levels of viremia upon the resumption of ART. The time needed to control viraemia was substantially reduced to 117 days after the first ATI and to 66 days after the second ATI.

The historical data from the primary infection for the other patients were unavailable, nonetheless we compared the highest VL attained, and the number of days taken by each patient to reach the undetectable VL after the second ATI. As shown in Figure 3, during the second ATI, patients exhibited approximately 16-fold reduction in the highest VL attained (Figure 3A, 3B) and a substantial 2 to 6-fold reduction in the number of days (median reduction by 53 days (range: 27 to 72 days)) required to return to undetectable levels (Figure 3C). These findings suggest that AGT103-T treatment enhances the immune system’s function, potentially enabling other therapies such as ART to act more rapidly and effectively. Further studies are warranted to determine if dose reduction or less frequent ART could be used in conjunction with AGT103-T to improve existing treatments for viral control.

**Figure 3:**
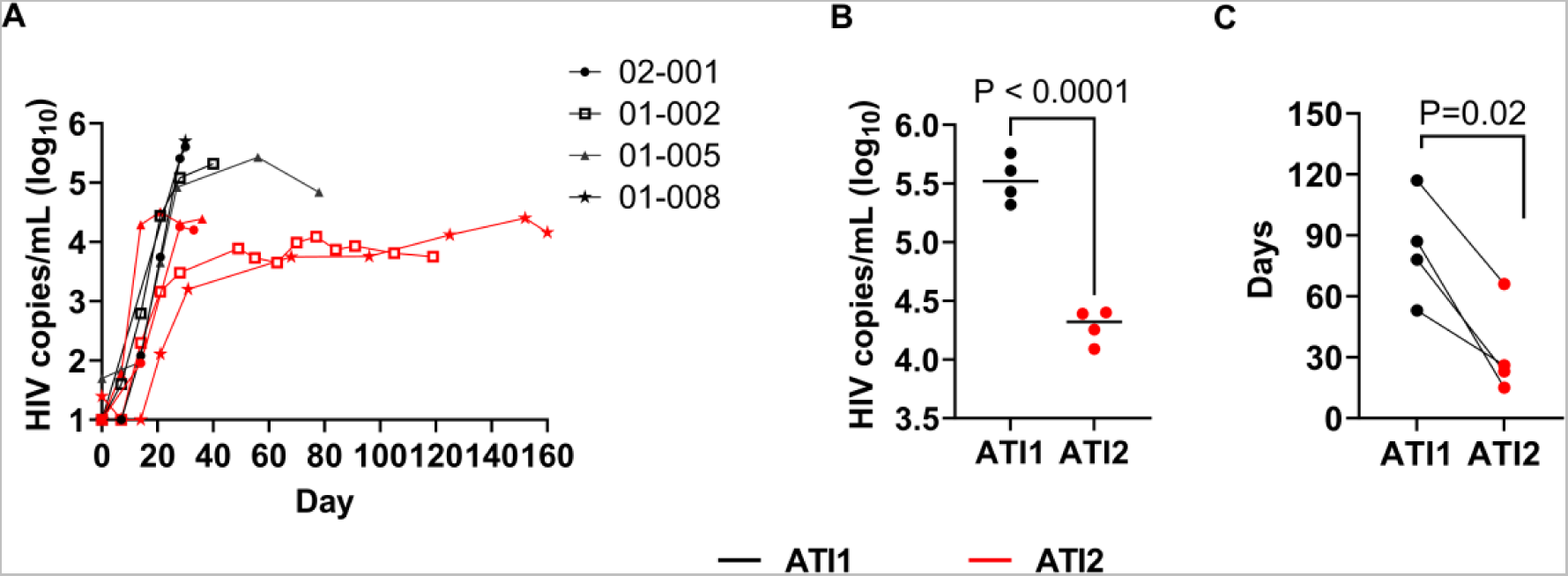
Impact of Multiple ATIs and AGT103-T Treatment on Viral Suppression. (A) Viral rebound during first (black lines) and second ATI (red lines) in four patients, 01-002, 01-005, 01-008 and 02-001. Each participant’s viral load (Log_10_HIV copies/mL) trajectory from the two ATIs are superimposed to provide a common temporal view of the viral rebound from time (days, x-axis) of each ATI initiation. (B) Maximum peak viremia attained by each participant during the first (black) and second (red) ATI before resuming ART (P <0.0001;). The black bar in each dataset represents the median of the four peak viremia values during each ATI. (C) Comparison of number of days required by each participant to reach ≤20 copies/mL levels of viremia after the first ATI (black) and the second ATI (red) (P=0.02). Student t-test (two-tailed) was performed to calculate the p-value.

## DISCUSSION

HIV remains a significant global health challenge, demanding continuous progress in therapeutic approaches. Decades of intensive research, particularly the discovery of delta Δ32 homozygous mutation in *CCR5* that led to a long-term remission from HIV has allowed the scientific community to develop strategies aimed at achieving HIV cure or a long-term HIV remission without the need for ART. Several experimental therapies including therapeutic vaccines, genetically modified immune cells that are resistant to HIV infection, cure inducing immunotherapies, interventions to permanently silence HIV infected cells, and drugs that reactivate latent viral reservoirs that can eventually be eradicated by CTL are being tested in the clinic. While no single therapy has offered a functional cure, it is highly possible that a combination of one or multitude of therapies might be required in the pursuit of HIV eradication and a functional cure.

In this study, we delve into the impact of AGT103-T, an autologous T-cell, gene therapy product, on CD8 T cell responses and viral suppression during multiple ATIs in individuals living with HIV. While the benefit of multiple ATIs in boosting immunity above the pre-therapy levels is controversial (2, 17, 20), we hypothesized that the presence of our drug product constituting HIV resistant Gag specific-CD4 T cells can enhance the magnitude of the CD8 T cell response, which, in-turn, can result in viral suppression. Since the original study (NCT04561258) was targeted to demonstrate the safety of AGT103-T in humans, it was not designed to integrate an ATI. However, one study subject discontinued their medication before the completion of the study, and it was possible that more patients would discontinue their ART. To address this and avoid unsupervised ATI, we developed a sponsor-initiated ATI study, NCT05540964, that aimed at assessing the impact of AGT103-T on the viral rebound, HIV immune responses, patient safety, and prevent the loss of critical information, in case the subjects ceased their ART without medical supervision. However, the ATI in the subjects was initiated well after infusion of the modified cells and the observed expansion of Gag specific CD4 T cells, 2-weeks post-infusion noted previously (12). The duration between the infusion and the ATI as well as the dose of the product was found to correlate with the persisting levels of AGT103-T in circulation (Figures 2, S1 and Table S1). Although the highest number of cells were detected in the patients who received the high dose of transduced cells, low number of persisting AGT103-T were also detected in all the other participants, irrespective of the dose exceeding two years post-infusion (Figures 2, S1). The levels of AGT103-T cells appeared to be modulated by the presence of viremia (Figures 2, S1), suggesting their engraftment and persistence as memory cells that can be re-activated with viral stimulation. In the absence of the viral pressure post-infusion, it is likely that a majority of AGT103-modified cells were lost which explains a sub-optimal immune response during the first exposure to viremia. The decline in the counts of AGT103-T cells with the rise in viremia as well as their partial restoration during ART, is also consistent with their migratory potential to the site of viral replication (25) although infection and depletion cannot be ruled out. Future studies are warranted to examine if these cells can be detected at other sites where feasible and if a concerted infusion of AGT103-T along with the ATIs are beneficial in generating immune responses capable of viral clearance.

Upon discontinuation of HAART during the first ATI, as anticipated, all participants experienced a resurgence of the virus, with a median VL of Log_10_ 5.4 copies/mL appearing within 30-56 days. Interestingly, one participant (01-007) achieved remarkable viral control for approximately six months (including a pre-enrollment patient-communicated ATI of 3 months) after discontinuing ART, underscoring the potential of AGT103-T in curtailing viral replication (Figure 1). This patient was newly diagnosed with HIV prior to commencing ART and studies have shown this group has a higher potential for establishing viral control after ATI (26, 27) and could represent a good target for future studies with larger numbers to determine an impact of AGT103-T on long term viral control. Contrary to other studies that employ ATI, our study noted a substantial increase in total CD8 T cell counts in 100% of the participants following the viral rebound. This is consistent with findings in a small population in other gene therapy studies (14, 18). During the second ATI, participants experienced a trend for a delayed viral rebound, with a significant decrease in peak viremia compared to the first ATI. The second ATI also revealed improved immune parameters, including a concurrent increase in CD8 T cell count and a potential role of Gag-specific CD4 T cells in enabling a Gag-specific CD8 T cell response, which could be responsible for the observed viral suppression. Upon resumption of ART, despite a modest decrease in the CD8 T cell counts observed, their overall levels remained elevated over baseline. This response is typically observed after the primary infection and can be attributed to persisting levels of antigen that causes immune activation as well as elevated levels of exhausted CD8 T cells (28). Additionally, an increased expression of Toll-like receptor 7 (TLR7) on CD8 T cells from HIV+ patients have also been shown to contribute to abnormal immune activation (29, 30). This poses questions as to what role AGT103-T could play if similar mechanisms are responsible for this observation. Overall, this suggests that AGT103-T treatment may contribute to a resetting of the immune response to that generated during primary infection.

Contrary to the theory of “auto-vaccination”, multiple ATIs are not always associated with viral suppression (17, 20). While some studies have observed a small suppression of post ATI viral set-point as compared to the pre-treatment VL of 0.4 Log_10_ copies/mL (20), others have reported no significant change in the magnitude of viral rebound (16, 17). Occasionally, a difference of 1 log_10_ copies/mL has been reported in studies with a small sample size (2). In our study, we had only one patient where pre-treatment VL was known and this patient, 01-005 demonstrated a 400-fold (2.6 Log_10_ copies/mL) reduction post-ATI rebound as compared to his pre-treatment viremia. Furthermore, all four study participants exhibited a difference of at least 1 log_10_ unit between the peak viremia during the first ATI and the viral set-point attained after the second ATI. A concomitant increase in immune response and a lower viral rebound, as a result of two ATIs, appears to be beneficial in preventing immune exhaustion whilst generating a sustained response necessary for viral suppression.

The significance of AGT103-T cells in generating an amplified systemic CTL response was particularly evident during the second ATI, although the patient size was very small (Figure 1). There was no correlation between the number of modified cells detected and the ability to suppress virus during the first ATI (Figures 2, S1). Furthermore, despite the presence of significant number of AGT103-T cells in both 01-008 and 02-009, both had similar viral rebounds during the first ATI to other patients. After the first viral exposure, 01-008 showed a concomitant CD8 response that led to viral suppression during the second ATI, underscoring the significance of at least two ATIs in the study design to evaluate the efficacy of a cell and gene therapy product to achieve viral suppression. Additionally, among the participants who underwent two ATIs, only two participants, 01-008 and 01-002, who demonstrated the presence of AGT103-T in their circulation at the start of the first ATI, were able to generate an amplified systemic CD8 T cell response during the second ATI. Since the Gag specific CD4 helper cells are critical for maintaining optimal CD8 T cell response as well as generating the memory T cells for future viral exposures, it is evident from our data that the two patients with higher numbers of AGT103-T cells during the first ATI, were able to generate a higher CD8 T cell response during the second ATI. Nevertheless, both participants observed a decline in the modified cells to very low levels with the rise in viremia, which might have impeded the generation of a sustained immune response. This suggests that while AGT103-T can help in generating an amplified CD8 T cell response, a therapeutic level of the cells in the presence of viremia might be critical to generate an optimal response capable of viral clearance. Thus, an ATI closer to the date of infusion coordinated with the time of expansion of the infused cells, might be beneficial in eliciting an immune response that is capable of ultimately clearing the virus.

While encouraging data was observed in this study, the small sample size precludes us from drawing many conclusions. As shown previously, a gradual decrease for both the Gag-specific cells as well as AGT103-modified cells were observed during the first six months after infusion of AGT103-T, resulting in marginal number of cells left in the circulation of all the participants. Despite the small number of AGT103-T cells detected in most patients, the observed viral suppression indicates the potential role of AGT103-T. Given that AGT103-T can suppress viral transcription by expressing siRNAs against HIV *Vif* and *Tat*, it is tempting to speculate that AGT103-T could have contributed to reduction in viral reservoir in the participants prior to their ART withdrawal. Since the standard primers used to detect viral reservoir in the intact proviral DNA assay (IPDA) also bind the lentiviral vector DNA, the proviral DNA levels in our subjects was not evaluated in this study, however, further studies should incorporate this as a crucial investigation to determine the efficacy of therapy. Nevertheless, the persistence of AGT103-T cells demonstrated resistance to viral clearance and responsiveness to immunological signals, offering a promising avenue for future HIV treatments. Overall, all participants exhibited improved viremia control after the second ATI and a faster response to ART. This implies that AGT103-T treatment enhances the immune system’s function, allowing other therapies like ART to act more swiftly in controlling the virus and limiting the viral reservoir. Crucially, all participants managed to achieve undetectable levels of viremia without encountering drug resistance or viral escape, and no HIV transmission was reported by participants, indicating a positive safety profile of AGT103-T in conjunction with two ATIs. Overall, our study underscores the potential of AGT103-T in enhancing CD8 T cell responses and viral suppression in individuals living with HIV. The observed immune enhancements, the “auto-vaccination” concept, and the persistence of AGT103-T cells present promising directions for future HIV treatment strategies. Further research incorporating multiple ATIs commenced closer to the time of infusion is essential to validate these findings and explore the full potential of AGT103-T in HIV therapy.

## METHODS

### Study Design

NCT05540964, the ATI study is a multi-center, safety, and efficacy study involving HIV positive participants who were previously infused with AGT103-T during the Phase 1 parent clinical study (NCT04561258). No investigational drug product was administered in this study. This study was designed for informational purposes to assess the impact of AGT103-T on host’s capacity to suppress HIV replication in the absence of ART, evaluate product parameters related to durable virus suppression, and assess participant immune characteristics related to durable virus suppression in those previously infused with AGT103-T. The trial commenced after obtaining institutional review board (IRB) approval and participant informed consent.

A baseline visit was conducted to confirm participant eligibility and establish baseline readings. Hematology, chemistry, coagulation, urinalysis, serum pregnancy, urine drug screen, hepatitis B and C screen, HIV viral load, CD4 T Cell Count, and CD4/CD8 ratio laboratory assessments were performed and recorded (LabCorp). After completion of the Screening and Baseline Visits, participants-initiated ATI, during which ART was withdrawn (Day 0). Participants who were taking non-nucleoside reverse transcriptase inhibitors (NNRTIs) were required to discontinue the NNRTI therapy 7 days prior to discontinuing the remaining therapies in the regimen. For these individuals, Day 0 was recorded as the day of discontinuation of all antiretroviral medications. Following ART discontinuation, participants were closely monitored for viral load, CD4, CD8 and lymphocyte count.

This study design comprised of two ATI phases. The dispositive studies were regular VL and a CD4 count determination. The patients were followed weekly for a month and biweekly thereafter until they met their ART resumption criteria. Therapy resumption was required if any VL exceeded >100,000 copies/mL of plasma in two successive visits or if the CD4 levels decreased below 300 cells/µL. Additionally, any unresolved study-related SAE or AE of grade 3 or greater could also lead to ART resumption. Following the conclusion of the first ATI, patients resumed their ART and were subsequently monitored at least monthly until VL value reached below 50 copies per mL and CD4 counts were recorded within 10% of baseline measurement. Once these baseline measurements were achieved, patients were offered the option to undergo a second ATI, with the same ART resumption criteria. Out of the six participants, two (01-007 and 02-009) withdrew before the start of the second ATI. Upon the resumption of ART after the second ATI, patients were followed biweekly until viremia was undetectable or for a maximum of 3 months after ART restart to document HIV VL suppression and assess CD4 recovery. Samples for secondary and exploratory endpoints were also collected at every visit.

In this study, the effects of the engrafted modified T cells (AGT103-T) on the kinetics of HIV rebound, duration of infection suppression, and measures of CD4 and CD8 T cell counts were assessed at time points specified in the schedule of assessments. Plasma viremia and CD4/CD8 T cell counts were also tested at every visit to monitor for indications that restarting ART might be advisable. To assess for genotypic drug resistance, a Monogram GenoSure PRIme test (LabCorp) was performed if the VL reached above 500 copies/mL to assure optimal anti-retroviral therapy. Additionally, the protocol included a provision that allowed for a participant to immediately restart ART if, at any point, the research volunteer, primary care provider, Principal Investigator, or Sponsor became concerned about the rising VL or the declining CD4 T cell count.

### Participants

Seven out of seven participants administered with AGT103-T in the parent study (NCT04561258) enrolled in the ATI study, however, participant 01-006 decided not to withdraw their ART. The participant information has been previously described in detail (12). Two out of the six participants (01-007 and 01-008) self-initiated their ATI prior to their official enrollment. Based on site-patient communication, 01-007 discontinued their ART after completion of the NCT04561258 study and remained viral rebound-free for 77 days prior to their enrollment. This claim was supported through an antiretroviral (ARV) drug test performed at the baseline visit, where no ARV was detected at the time of enrollment. Due to personal reasons, 01-007 withdrew from the study before its conclusion. In contrast, 01-008 had self-initiated ATI during the parent study (NCT04561258), approximately 150 days post-infusion, and was found to have 570,000 viral copies/mL on their day 180 visit. Thus, the participant was promptly advised to resume their ART medication. Once the participant achieved an undetectable viral load, the participant was invited to enroll in the ATI study and initiate their second ATI.

### Assessments of safety

Adverse events were graded following the National Cancer Institute’s Common Terminology Criteria for Adverse Events, version 5 (CTCAE, v5).

### Intracellular cytokine staining (ICS) assay for detecting HIV-specific CD4 and CD8 T cells

The ICS assay was performed as described previously (12). Briefly, PBMCs were isolated from whole blood using Ficoll Plaque and Sepmate Separation tubes (StemCell Technologies) per manufacturer’s instructions. These PBMCs were then cryopreserved in RPMI (ThermoFisher Scientific) containing 10% human AB serum (hABS, MilliporeSigma) and 7% DMSO (MilliporeSigma) until ready for analysis. Cryopreserved PBMC vials were viably thawed, and cells were cultured for 18–22 h in RPMI + 5% hABS. The following day, cells were counted and resuspended in fresh RPMI + 5% hABS and stimulated with 1 µg/mL pooled overlapping peptides spanning HIV Gag (PepMix HIV-1 GAG Ultra; JPT Peptide Technologies), in the presence of GolgiPlug (1:1000; BD Biosciences) for 4 h at 37°C and 5% CO_2_. Medium containing 0.4% DMSO (peptide resuspension media) served as a negative control, and CEF/CPI Class I/II-restricted peptides (Cellular Technology Limited) were used as positive controls for responses to common antigens. After stimulation, cells were collected and washed in Dulbecco’s phosphate-buffered saline (DPBS, ThermoFisher Scientific) and stained with Fixable Viability Stain 450 (BD Biosciences) to exclude dead cells. Subsequently, cells were washed in FACS staining buffer, blocked in TruStain Fc blocking solution (Biolegend) and then stained with anti CD3-PerCP, CD4-FITC, andCD8-PE (Biolegend) antibodies, followed by fixation using 4% paraformaldehyde (EMS). Fixed cells were permeabilized in 1X perm/wash buffer (BD Biosciences) followed by intracellular staining with IFNγ-APC antibody (Biolegend). After washing with perm/wash buffer, cells were stored in FACS staining buffer until ready for analysis using a BD FACSLyric flow cytometer. Data was analyzed using Flowjo software (BD Biosciences). To count the number of CD4 and CD8 T cells expressing IFNγ, the gating strategy was described previously (12). Results were expressed as HIV Gag-specific CD4+ and CD8+ T cell count per mL of blood.

### Detection of AGT103-modified T cells by q-PCR based vector copy number (VCN) assay

The assays for detecting the frequency of transgene-containing AGT103-T cells was adapted from our previous study (12) with some modifications that was shown to enhance the sensitivity by about 4-fold enabling the detection of AGT103+ cells at levels as low as 0.001% in total circulating PBMCs. Briefly, whole blood was collected during visits in K2 EDTA tubes (BD Biosciences), which were then processed by Ficoll (GE) separation to isolate PBMC. One vial with 10 million frozen PBMCs was then thawed, and the cell viability was determined. These cells were then used to isolate an enriched CD4 T cell population using REALease® CD4 MicroBead Kit according to manufacturer’s instructions (Miltenyi). Total DNA was extracted from CD4 T cell pellets (approximately 0.2 to 1 million cells) using Dneasy DNA extraction kit (Qiagen) to determine transgene copy number by TaqMan (Thermofisher Scientific) quantitative polymerase chain reaction (qPCR), targeting WPRE sequence, a common feature of lentiviral vectors including AGT103. Copy number was calculated by normalizing the copies of WPRE detected with a single-copy housekeeping gene, *RPL32* (24). The following primer and probe sequences were used to amplify and detect the respective gene sequences. WPRE FP: 5’-GCTATGTGGATACGCTGCTTTA-3’, WPRE RP: 5’-AGAGACAGCAACCAGGATTTATAC-3’, WPRE probe 5’-/56-FAM/TCATGCTAT/ZEN/TGCTTCCCGTATGGCT/3IABkFQ/-3’, RPL32 FP:5’-CAAGGAAAGACGAGCTGTAGG-3’, RPL32 RP:5’-GGGCAGTTGCATCTTCATATTC-3’, RPL32 probe 5’-/5HEX/AGCTGCAGG/ZEN/CAGAAATTCTGGTAGT/3IABkFQ/-3’. Given that the vector copy number (VCN) per transduced cell in all the drug products infused into these patients was less than one, the lentiviral copies per cell were converted into the number of AGT103-T cells. Using the CD4 T cell count obtained from LabCorp, the lentiviral copies per cell or the number of AGT103-T cells were then expressed as either a proportion of circulating CD4 T cells or per mL of blood.

### Modified miRNA-CCR5 detection by amplicon sequencing

To detect the presence of modified miRNA targeting CCR5, we employed the Illumina Miseq platform and the 2x KAPA HiFi HotStart ReadyMix (Roche) kit for amplicon sequencing, performed at Psomagen, Rockville, MD. The process was followed according to the manufacturer’s instructions. In brief, total DNA was extracted from purified CD4 T cell pellets, consisting of approximately 0.2 to 1 million cells. The extracted DNA underwent a meticulous assessment for both quality and quantity using DNA electrophoresis and fluorometric measurements. Subsequently, PCR amplification step was conducted to amplify the miR30 sequences flanking the site of insertion of shRNACCR5 in our LVV using the following primers: AdapmiRNA30F TCGTCGGCAGCGTCAGATGTGTATAAGAGACAGAGGTATATTGCTGTTGACAGTGAGCGACTG TA and >AdapmiRNA30R GTCTCGTGGGCTCGGAGATGTGTATAAGAGACAGAGCCCCTTGAAGTCCGAGGCAGTAG. The PCR reactions adhered to cycling conditions that had been rigorously optimized for the amplification of these sequences. The resulting amplicon libraries underwent further evaluation for quality and quantity using TapeStation D5000 Screen Tape and DNA QC-Picogreen assays. For the sequencing step, we performed paired-end sequencing on the Illumina Miseq platform, generating 151 base pair reads. The sequencing process followed Illumina’s standard protocols, ensuring data integrity, and minimizing bias. The sequence data generated from the Illumina platform were then subjected to comprehensive processing and analysis using specialized bioinformatics tools. We employed bioinformatic pipelines to identify and quantify the presence of human endogenous miR30 (GRCh38 chr6: 71403551-71403621) and modified miR30 targeting *CCR5* within the samples. Given that full miR30 is present as a single copy gene in the genome, the ratio of modified miR30 to native miR30 was used to determine the proportion of AGT103-containing cells among the circulating CD4 T cells.

### Statistical evaluation

All calculations were performed using Microsoft Excel. P-values to determine statistical significance were obtained through a Student’s t-test (two-tailed). Gag-specific T cell count was calculated based on the percentages obtained in FlowJo™ v10.8 Software (BD Life Sciences) using the following equation. % of IFNγ+ CD4 (or CD8)/100 * number of CD4 (or CD8)/µL blood * 1000µL (1mL) = total Gag Specific CD4 (or CD8)/mL blood. The number of AGT103-T cells per mL was calculated by multiplying the vector copy number (VCN) per cell by the total CD4 T cell count per mL. To express the number of AGT103-T cells as a proportion or percentage of circulating CD4 T cells, the VCN per cell was multiplied by 100. The viral set-point was determined by calculating the average of the VL readings obtained after the exponential increase in viremia reached a plateau.

### Study Approval

This clinical trial was approved by the central IRB of Advarra, Inc. and the IRB of Georgetown University Medical School. Written informed consent was obtained from all the participants prior to their enrollment on the clinical trial. The clinical trial is registered at Clinicaltrials.gov (NCT05540964)

## Data availability

The datasets presented in this article are property of American Gene Technologies International, Inc. Requests to access datasets should be directed to the corresponding author.

## Author Contribution

AJ and JSB wrote the manuscript. AJ, GEC and JSB edited the manuscript. AJ, GEC, ML, AIC, LX, IM and JSB contributed to the development of the assays and analyzed the data. GEC, ML, AIC, LX and IM performed the experiments. ELF and MAC managed the clinical and regulatory operations of the study. JAG provided the funding. PNK and JB contributed to the development of the clinical trial protocol, recruiting, treating, and collecting blood samples from the participants. MAC and JSB conceived, designed, and supervised the study. All authors contributed to data interpretation and reviewed the final manuscript.

## Supporting information

Sipplemental figure 1 and Supplemental Table 1

## Data Availability

All data produced in the present study are available upon reasonable request to the authors.

