## Supplementary material for "Multiple Treatment Interruptions and Protecting HIV-Specific CD4 T-Cells Enables Durable CTL Response and Viral Control": Sipplemental figure 1 and Supplemental Table 1

Supplementary data

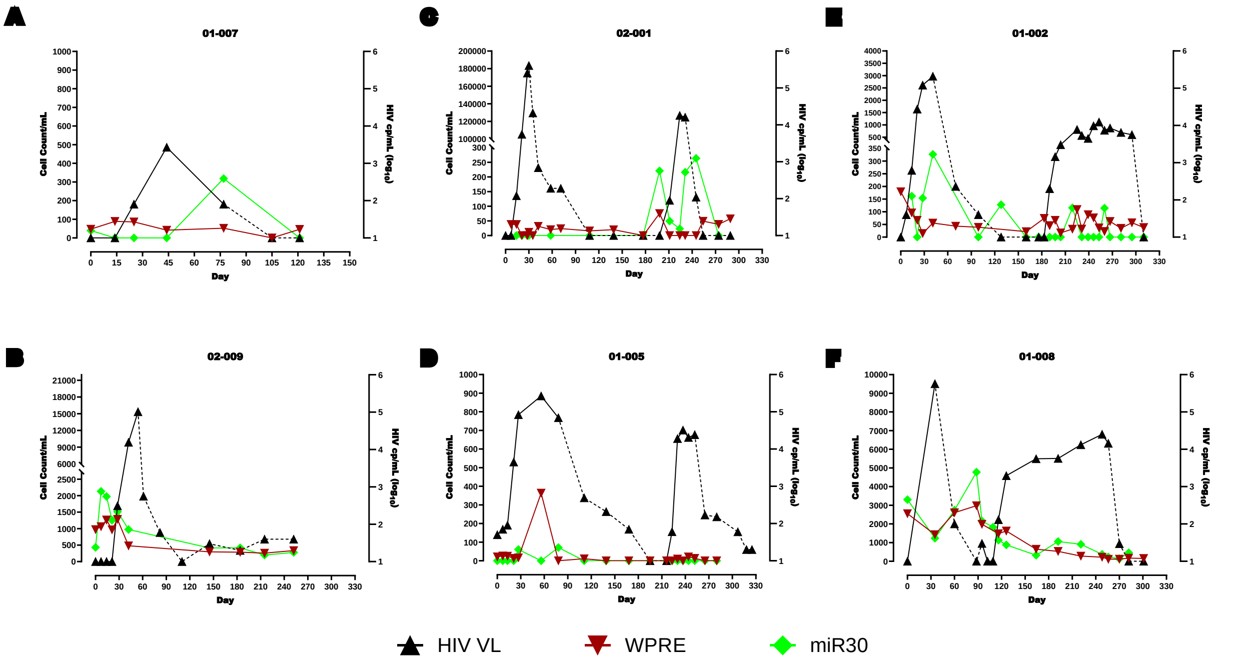


**Figure S1: Comparison of AGT103-T cells detected by two independent methods.** (A-F) AGT103-T cells detected at various timepoints during the study by two independent methods, qPCR and amplicon-sequencing in six study participants were plotted against the number of days (x-axis) and the viral load (black). The q-PCR method detected WPRE normalized against housekeeping RPL32 (red) and amplicon-seq detected the ratio of modified miR30 to native miR30 (green). The cell counts are plotted on the left-hand Y-axis while the VL is reported on the right-hand Y-axis. The solid black line indicates the VL during the ATI and the dashed black line represents the VL on ART.

**Table S1: AGT103-T cells as a proportion of circulating CD4 T cells relative to the dose and duration between the infusion and the initiation of the ATI.**

| Patient ID | Infused Product Dose | Days between Infusion and the Start of ATI-1 | AGT103-T (% of total CD4) at the Start of ATI-1 |
|---|---|---|---|
| 01-008 | 1.67 E+9 | 150 | 0.544 |
| 02-009 | 1.38 E+9 | 99 | 0.220 |
| 01-002 | 0.192 E+9 | 490 | 0.026 |
| 02-001 | 0.62 E+9 | 246 | 0.008 |
| 01-005 | 0.46 E+9 | 411 | 0.003 |
| 01-007 | 0.19 E+9 | 390 | 0.009 |
